# Tissue expression in surgically retrieved cam deformity and capsule from patient hips with Cam-type Femoroacetabular Impingement Syndrome

**DOI:** 10.1101/2024.07.02.24309871

**Authors:** Catherine Yuh, Philip Malloy, Steven P Mell, Zeeshan Khan, Shane J. Nho, Robin Pourzal, Jorge Chahla, Deborah J Hall

## Abstract

**Introduction:** Cam-type femoroacetabular impingement syndrome (FAIS) is a pre-arthritic hip condition, defined as a bony growth on the proximal femur that causes abnormal joint contact. The tissue presentation of the cam deformity and capsule in FAIS remains understudied. The purpose of this study was to 1) evaluate histopathological features in cam deformity and capsule from FAIS patients, 2) assess the extent of local inflammation within the capsule, and 3) determine relationships between cam deformity tissue composition versus α angle and patient factors.

**Methods:** Cam deformity and capsular tissues were collected from FAIS patients undergoing arthroscopic surgery. Samples were histologically processed, imaged using light and polarized light microscopy, and assessed with point counting. Correlation-based statistics were performed to identify features correlated with α angle and patient factors.

**Results:** Across 21 cam deformity samples assessed, a total of 16,259 points were counted. The tissue within the cam deformity was observed to be heterogeneous between specimens, comprised of 16 distinct structures spanning different states of viability. In samples with articular cartilage, the tissue was highly disrupted and/or calcified. The presence of fibrocartilage, necrotic cartilage, and vasculature had significant low-moderate correlations with α angle. During assessment of capsular tissue quality, synovitis was observed in most samples.

**Conclusion:** The cam deformity is complex and heterogeneous, both within individual cam deformities and between individuals with FAIS. Several cam deformity tissue features were correlated with α angle, age, sex, and BMI. The heterogeneity observed in these samples indicates that tissue properties within the cam deformity varies between patients with FAIS, which may contribute to outcomes of hip arthroscopic surgery and a patient’s level of risk for the subsequent development of osteoarthritis. Our findings suggest distinct tissue phenotypes of FAIS exist, which may be an important consideration for FAIS treatment strategies.

## INTRODUCTION

Femoroacetabular impingement syndrome (FAIS) is a hip disorder that is especially common in active individuals and high-performance athletes, with a reported prevalence of up to 89%^1,2^. A common cause of FAIS is cam morphology, characterized as a ‘bony’ growth that causes an asphericity of the femoral head^3^, leading to a lack of femoral head-neck offset. Cam morphology is widely considered a pre-arthritic condition, acting as a precursor for hip osteoarthritis (OA)^3–7^. The positive predictive value of developing hip OA within 5 years is estimated to be 10.9 and 25% in patients diagnosed with moderate and severe cam morphology, respectively^3^.

Since the formal conceptualization of FAIS as a distinctive disease by Ganz et al.^5^, most studies have been clinically-focused, with an emphasis on advancing surgical techniques to improve clinical outcomes^8–10^. Hip arthroscopic surgery is the primary treatment for FAIS to reduce pain and restore function^11–13^ with excellent short to midterm patient-reported outcomes^10,11^. However, despite these promising outcomes, multiple factors have been reported to be predictive of poor outcomes including older age, female sex, higher body mass index (BMI), and larger size of cam deformity^10,14^. Open and arthroscopic surgical techniques for FAIS were designed based on the sole premise that removing the cam deformity would improve hip biomechanics. However, it remains unknown if the underlying tissue pathology associated with FAIS is resolved by elimination of the mechanical impingement and/or related to the subsequent development of hip OA. Thus, studying the tissue properties associated with FAIS can provide crucial insight to guide future targeted treatments.

While studies have reported on articular cartilage presentation with FAIS^15–17^, studies to robustly or quantitatively investigate the tissue-level structures and viability of the cam deformity are limited. Recently, Youngman et al.^18^ reported the detection of non-osseous and soft tissues at the cam deformity using magnetic resonance imaging (MRI), demonstrating that heterogeneities may exist at the tissue-level between different cam deformities. It remains unknown whether tissue-level heterogeneity can be attributed to disease severity, OA progression, or other factors. This uncertainty presents a barrier to understanding multiple aspects of FAIS, including disease progression, and may be a potential contributing factor towards unfavorable surgical outcomes in certain individuals. Thus, investigations into the relationship between the tissue properties versus disease severity and patient factors may aid in further understanding the progression of FAIS disease and identifying potential features that may be predictive of subsequent OA development.

Another tissue system that contributes to inflammation and pain in musculoskeletal conditions, such as OA, is the surrounding capsule tissue ^19,20^. While studies^21,22^ have assessed joint-level metrics of capsule morphology, such as MRI-derived dimensions, thickness and volume in the context of FAIS, there is limited research on capsular tissue morphology. Considering the capsule has been implicated as a source of local inflammation of the joint in OA^19,20^, assessing the extent of the capsular damage or inflammation may also provide insight into the progression of FAIS into OA.

Here, we investigated the cellular and tissue environment within cam deformity and capsule tissues from cam FAIS patients undergoing hip arthroscopy. Point counting was performed to spatially characterize the specimens based on tissue type and quality, allowing for quantitative and statistical assessment. The aims of this study were to: 1) Evaluate histopathological features in surgically-retrieved cam deformity and capsular tissues, from patients undergoing hip arthroscopy for cam-type FAIS, 2) Assess the extent of local inflammation within the joint capsule, and 3) Determine the relationship between the histopathological tissue composition of the cam deformity versus α angle and patient demographics.

## METHODS

### Subjects and Institutional Approval

An institutional review board protocol with an associated HIPAA waiver of consent was approved for this study. Only tissue that would be otherwise discarded during the normal course of surgery was procured. The duration of this study was one year, and a total number of N=23 samples were collected. Two samples were excluded due to a lack of sample volume, leading to artifacts during histological preparation.

### Surgical procurement

Inclusion criteria included the following: 1) patients >18 years of age, 2) clinical diagnosis of FAIS, 3) electing to undergo hip arthroscopy. Exclusion criteria included prior diagnosis of avascular necrosis, dysplasia, and previous history of hip surgery. Patients diagnosed with FAIS undergoing primary hip arthroscopy with a fellowship-trained sports medicine surgeon at a single institution were included. Diagnosis of FAIS was made based on reports of hip pain and symptoms, positive clinical exam findings and radiographic evidence of cam morphology. Alpha angles were measured using Dunn 45° hip x-ray views, by drawing a circle on the femoral head and measuring the angle between the line from the center of the femoral head to the midpoint of the femoral neck and another line from the center of the femoral head to the point of discontinuity in the circle.

Tissue samples were collected from the apex of the cam deformity and capsule of each subject. The operative joint was accessed via a standard anterolateral portal (ALP) under fluoroscopic guidance (Figure 1). A second modified mid-anterior portal (mMAP) was established. While visualizing the cam deformity arthroscopically from the ALP, an arthroscopic curette was passed through the mMAP and used to harvest cam deformity tissue prior to resection. Capsular tissue was collected using an arthroscopic punch scissor. Samples were immediately stored in 10% formalin.

**Figure 1:**
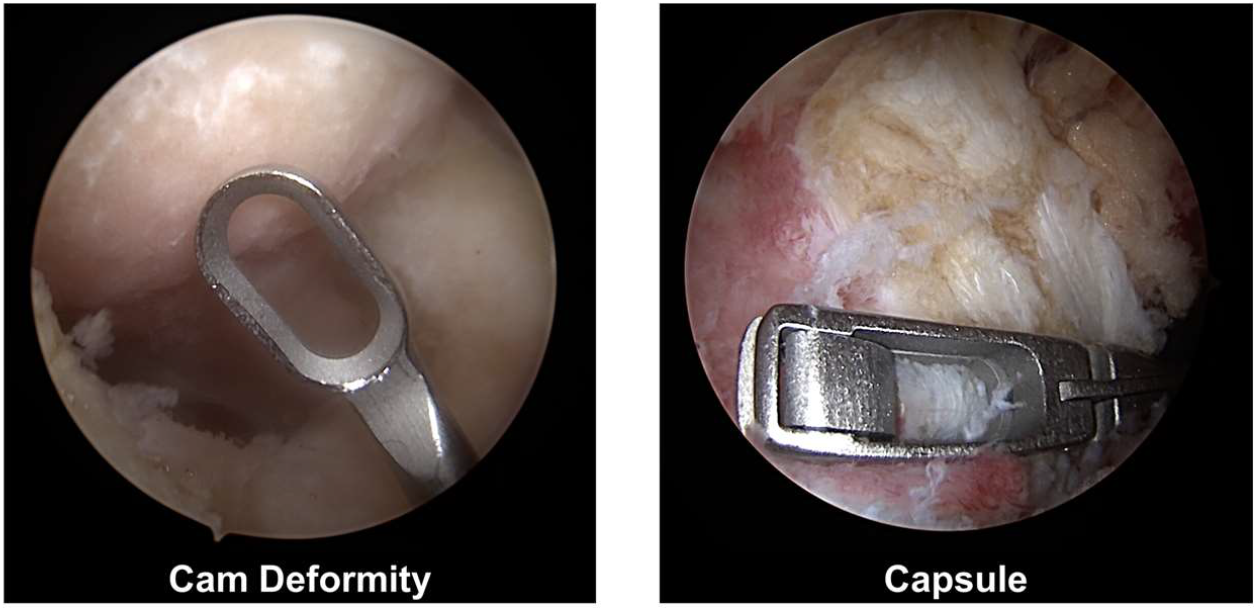
Arthroscopic images indicating areas where specimens were procured. From each subject, specimens were obtained from the apex of the cam lesion and the capsule.

### Histological preparation and assessment

All samples were processed for paraffin-embedding. For cam deformity samples, decalcification was performed due to the presence of bone. Samples were sectioned (6-μm) and stained with hematoxylin and eosin. Light microscopy (Eclipse 89*i*, Nikon, Melville, NY) was used to assess tissue morphology. Polarized microscopy (i.e., birefringence) was performed to assess the presence of fibrillar structures and to discern tissues that have characteristic birefringent signals, such as bone and fibrocartilage. Two experienced reviewers assessed the images together, using a consensus-based method. Discrepancies between the reviewers were discussed until consensus was made on the tissue classification.

Point counting was performed on cam deformity samples. A grid of lines 25 μm apart was overlayed on each sample (Figure 2). At each intersection, samples were carefully assessed for tissue structures and viability, the latter of which was defined by two criteria: exhibiting nucleated cells and exhibiting characteristic extracellular matrix structure with no disruption. A spatially-based percentage of the total specimen for each feature was then calculated. Across the 21 samples assessed, a total of 16,259 points were counted. Point counting was not implemented for the capsule, as they did not exhibit the tissue heterogeneity seen in the cam deformity. Instead, for capsular samples, a binary assessment was performed where each feature was marked ‘present’ or ‘not present’. For the capsule, tissue quality was assessed based on the presence and quality of the synovial lining, the orientation of the collagen fibrillar network based on birefringence signal, vascularity, cellular activity, and secondary features (bone fragments, inflammation, etc.).

**Figure 2:**
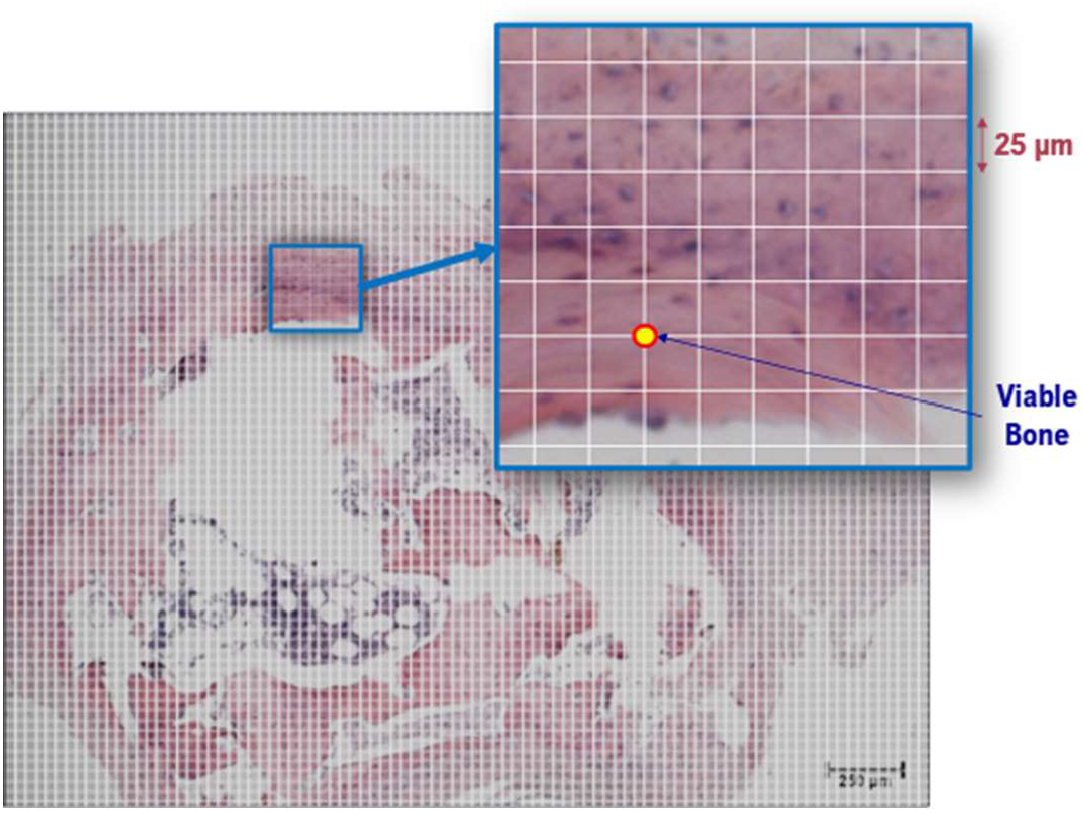
Representative image with point counting grid overlaid. A grid of lines, 25 μm apart, was defined, and the feature at each intersection was recorded.

### Statistical Analysis

Pearson’s correlation coefficients were calculated for each feature observed during point counting of the cam deformity samples to determine if the spatially-based percentage of each tissue feature was correlated to α angle, age, sex, and BMI. 95% confidence intervals were estimated with bootstrapping (random resampling with replacement) to improve overall statistical power and accuracy of our statistical estimates^23,24^. If the 95% confidence interval did not include 0, we rejected the null hypothesis that there is no correlation.

## RESULTS

### Cam Deformity Histopathological Features

Large variations of tissue structures and viability status were observed across the cam deformity samples (Figure 3). We identified 16 unique tissue features, across 8 different tissue types (Figure 4). As expected, bone was a prominent feature across most samples, although its state varied widely from viable, detritus, or necrotic. Interestingly, some samples exhibited minimal bone relative to the entire sample. Although the presence of cartilage was not as abundant compared to bone, several cartilage types were identified within the cam deformity, including hyaline cartilage, fibrocartilage, and calcified cartilage. In cam deformity samples that presented with cartilage, the cartilage appeared ‘abnormal’ (highly disrupted, fibrotic, or necrotic).

**Figure 3:**
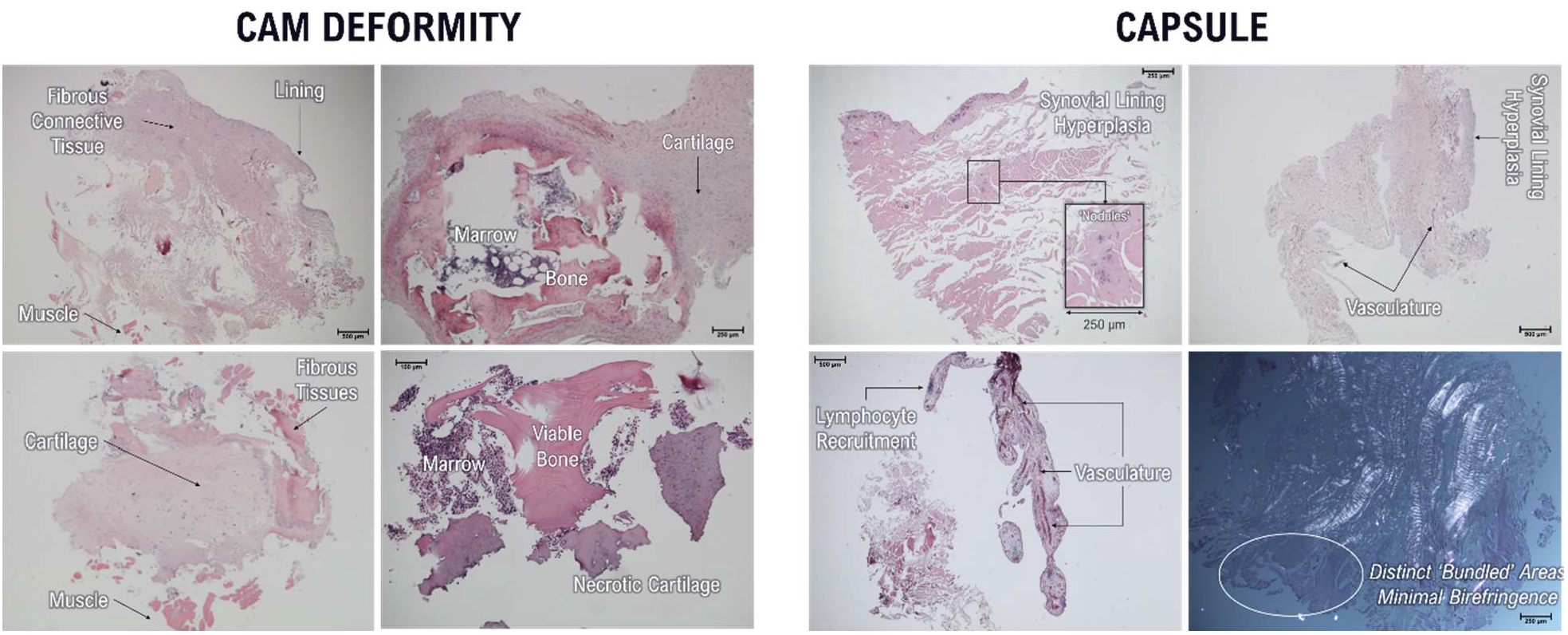
Left: Representative images of cam deformity specimens. Variations in tissue features can be seen across different specimens. Right: Representative synovium/capsular images from different patients.

**Figure 4:**
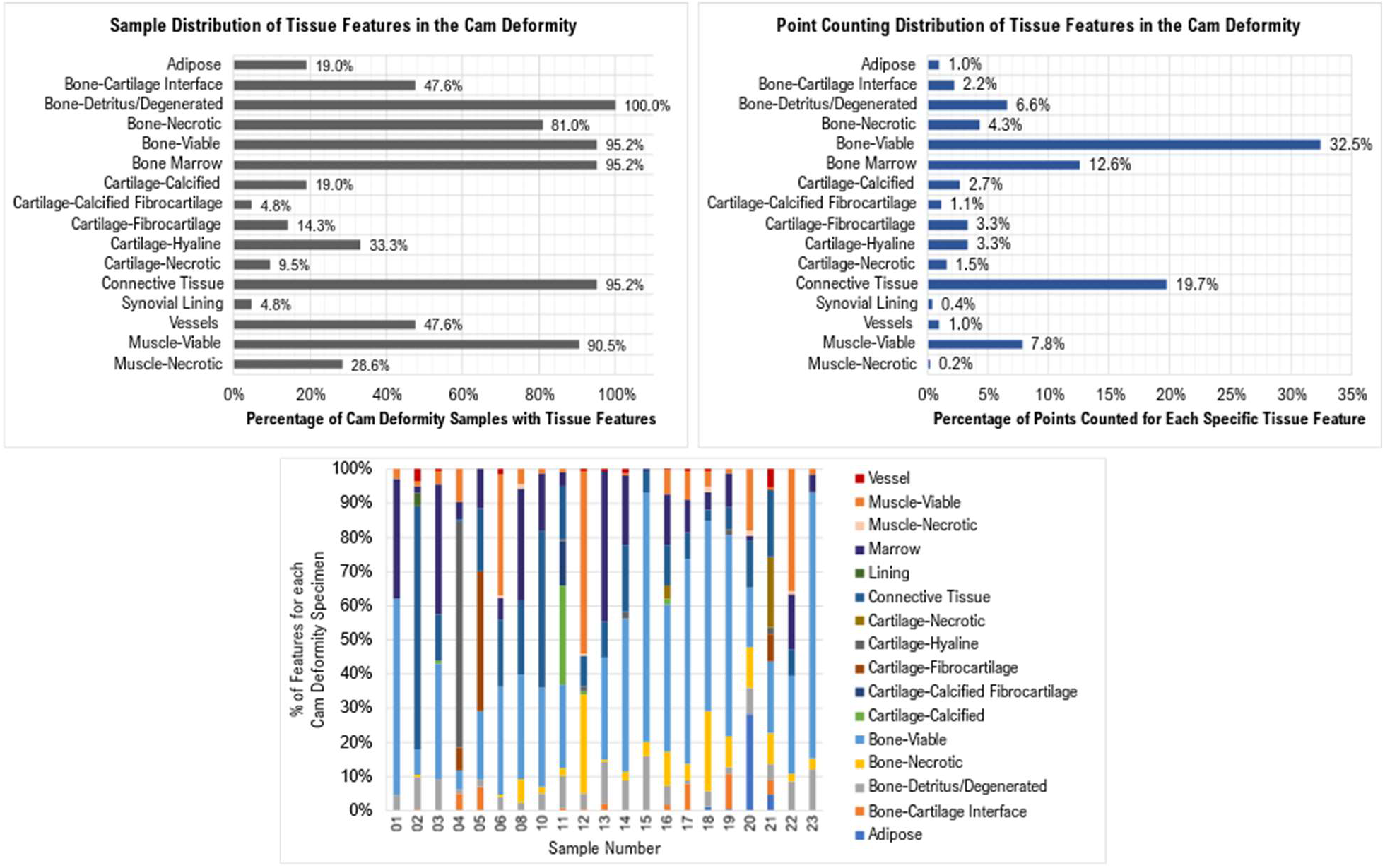
Overall distribution of tissue features in the cam deformity, determined from the binary assessment of whether tissue features were present or not. Top Left – Distribution based on the number of samples exhibiting each tissue feature. Top Right – Distribution based on the number of points counted exhibiting each tissue feature. Bottom – The percentage for each tissue feature shown within each specimen (represented by each bar).

Connective tissues were identified as the second most observed tissue feature in the cam deformity, with a 95.2% observation rate across all samples and a 19.72% observation rate based on points counted in the cam deformity (Figure 4). To properly describe these connective tissue structures, qualifiers were used to describe the density of the tissue network (loose/dense) and the presence of fibers (birefringence). Most samples exhibited loose connective tissue, with very few samples exhibiting dense connective tissues.

While muscle was present in 90.5% of specimens, only 7.8% of points counted were comprised of muscle, indicating that this was likely an artifact of port placement during arthroscopy.

### Capsule Histopathological Features and Local Inflammation

Capsular tissues were found to be more consistent in quality across samples, sharply contrasting our findings for the cam deformity. Interestingly, the joint capsule samples exhibited minimal cellular activity that would indicate chronic local inflammation – 2 out of 21 samples exhibited an inflammatory response, characterized by hypercellularity and the presence of specific immune cells, such as lymphocytes or plasma cells. While inflammatory cells were not abundantly observed across the capsule specimens, if at all, several abnormal features in the capsule were identified (Table 1). Out of 21 samples, 7 presented with hyperplasia and/or thickening of the synovial lining, often with hypervascularity, indicating potential synovitis. Other ‘abnormal’ features observed included the presence of ‘nodule-like’ tissue features (Figure 3). While these features closely resembled the structure of blood vessels, these were often closed with no indication of nearby red blood cells. Because of this lack of certainty, these features were designated to a separate tissue category. Another ‘abnormal feature’ identified in many samples were tissue structures that appeared ‘bundle-like’ that expressed minimal to no birefringence, appearing in 11 samples. In these areas, a clear demarcation of structures was present, both visible under bright light and polarized light. In addition, similar ‘bundled’ tissue structures with birefringence were also observed in 7 samples.

**Table 1:** Overview of various tissue features and markers of quality in the capsule tissue for each specimen.

| Sample | Lining | Cellularity | Vascularity | Bone Fragments | Bundles (Birefringence) | Bundles (No Birefringence) | Inflammation | Fascia/Adipose | Nodule |
| --- | --- | --- | --- | --- | --- | --- | --- | --- | --- |
| 1 | No Lining | Normal | Normal | X |  |  |  |  |  |
| 2 | Hyper | Normal | Hypervascularity |  | X |  |  |  |  |
| 3 | No Lining | Normal | Normal | X |  | X |  |  |  |
| 4 | No Lining | Normal | No Vessels |  |  | X |  |  |  |
| 5 | Hyper | Hyper | Hypervascularity |  |  |  | X |  |  |
| 6 | No Lining | Normal | Thickened vessel walls |  |  | X |  | X |  |
| 8 | No Lining | Hypo | Large Vessels |  |  | X |  | X |  |
| 10 | Hyper | Hyper | Hypervascularity | X | X |  | X |  | X |
| 11 | Hyper | Normal | Hypervascularity | X | X |  |  |  | X |
| 12 | No Lining | Normal | Normal | X |  | X |  |  | X |
| 13 | Hyper | Normal | Hypervascularity |  |  | X |  |  | X |
| 14 | No Lining | Normal | No Vessels | X |  |  |  |  |  |
| 15 | No Lining | Normal | No Vessels | X |  |  |  | X |  |
| 16 | Hyper | Normal | Normal |  |  | X |  |  |  |
| 17 | No Lining | Normal | Normal | X | X | X |  |  | X |
| 18 | Hyper | Normal | No Vessels |  | X |  |  |  | X |
| 19 | No Lining | Normal | Normal | X | X | X |  |  |  |
| 20 | Normal | Normal | Normal | X | X | X |  |  |  |
| 21 | No Lining | Normal | Normal |  |  | X |  |  |  |
| 22 | No Lining | Normal | Normal |  |  |  |  |  |  |
| 23 | Normal | Normal | Normal |  |  |  |  |  |  |

### Relationships Between Cam Deformity Tissue Features Versus Disease Severity and Patient Factors

Out of 16 unique tissue features observed in the cam deformity, the spatial percentages of fibrocartilage, necrotic cartilage, and vasculature were shown to have significant moderate positive correlations with α angle (Table 2). In terms of demographic and anthropometric factors, several tissue features were found to be significantly correlated to these factors (Table 2). Age was found to be negatively correlated to the percentage of detritus or degenerated bone, and positively correlated to the percentage of hyaline cartilage. Necrotic bone and viable muscle were both found to be moderately correlated to male sex (negative correlation). Fibrocartilage, connective tissue, and bone marrow presence were found to have low to moderate correlations to female sex (positive correlation). Finally, BMI was found to be negatively correlated with the percentage of hyaline cartilage and vasculature.

**Table 2:**
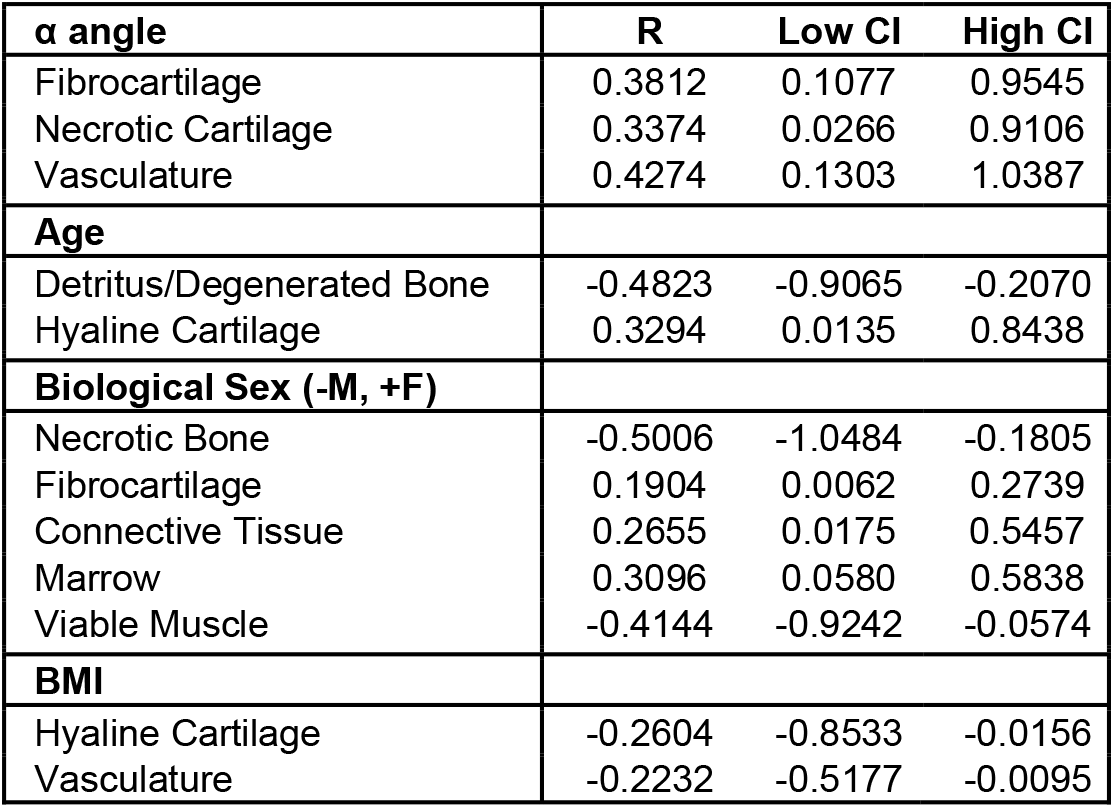
Pearson’s correlation coefficients with 95% confidence intervals estimated by bootstrapping for tissue features found to be correlated with disease severity, age, biological sex, and BMI.

## DISCUSSION

This study demonstrates that the cam deformity is comprised of a heterogeneous mixture of tissue types, including bone, cartilage, and fibrous tissues. The state of viability of these tissues also varied widely between subjects, with viable tissue, detritus tissue, and necrotic tissue observed across the samples. Only minimal inflammation was observed in respective capsular specimens. Additionally, certain features were found to be significantly correlated to α angle and various patient demographics. We also qualitatively observed differences in tissue structure distribution between patient samples, which may indicate that cam-type FAIS may present as different tissue phenotypes of disease, as opposed to a singular disease characterized simply by an osseous overgrowth. The notion of cam-type FAIS phenotypes has also been previously suggested in other literature^15,23,24^, and our results strongly support this paradigm.

To our knowledge, this is the first study to quantitatively assess the tissue features within the cam deformity from FAIS patients. While there are previous studies assessing the cam deformity and surrounding tissues, most studies have relied on cadavers, or have implemented histological grading scales^15,17,25,26^ of cartilage degradation that were specifically developed to describe OA progression. While these previous studies have offered crucial insight into tissue structures associated with FAIS, the tissue quality is often qualitatively described, limiting the ability to assess relationships between tissue expression and other factors. Here, we implemented point counting to determine a spatial percentage for each tissue feature, providing a robust quantification of presence of certain features in the cam deformity and a statistical assessment of the relationships between tissue features and α angle.

We identified 16 distinct features across 8 distinct tissue types within the cam deformity. Of these features, 3 were correlated with increasing α-angle: fibrocartilage, necrotic cartilage, and vasculature. These tissue features have all been reported to be associated with tissue injury and repair^27,28^. Given that FAIS induces a repetitive injury at the proximal femur, these potential relationships between cam deformity size (α angle) and tissue markers associated with injury are probable. Future work is necessary to determine if there is a mechanistic link between these factors and cam deformity tissue presentation and will include assessing injury markers using targeted techniques like immunohistochemistry and spatial transcriptomics. In addition, while capsular specimens exhibited minimal to no local inflammation, the presence of both hyperplasia and hypervascularity is suggestive of synovitis, which has been shown to be associated with pain^29^. Features that were uncharacteristic or abnormal, such as closed nodules of concentric cells or bundle-like tissues, were also identified. For the latter, it may be of interest to determine whether differences in fiber density of these bundle-like tissue structures are related to the tissue’s structural integrity. Future work is also necessary to determine what the nodule and bundle-like features represent and how they are related to capsule health/pathology.

In addition to disease severity, age, sex, and BMI were assessed. Given that FAIS typically occurs in younger individuals, it is reasonable to speculate that mechanisms associated with skeletal maturity may contribute to the tissue distribution observed in the cam deformity. This would correspond to our finding that age was negatively correlated with detritus bone, which may be due to increases in mature bone in older individuals. Interestingly, we did not observe the same correlations between age and viable bone-only detritus bone was found to be correlated with age. This suggests that skeletal maturity may not be the sole contributing factor to the tissue features observed, and that other factors, such as type of sports/daily activity, joint biomechanics, and anatomical variations may also be involved. In addition, hyaline cartilage was shown to have a positive correlation with age. This was contradictory to what we expected, since hyaline cartilage tends to be more abundant in younger developing individuals who are not yet skeletally mature. One possible explanation as to why hyaline cartilage was found to increase with age may be related to the changes in architecture associated with cartilage maturation. Compared to immature cartilage, mature cartilage has a hierarchal architecture that contributes to an increased mechanical stiffness and increased stability of the extracellular matrix network^30^. These characteristics are known to play roles in the tissue’s capacity to bear load^31–33^. This may have implications on how the tissue responds to abnormal loading incurred by repetitive joint injury, as occurs in FAIS. While this speculation may be reasonable, it is crucial to note that only a low-moderate correlation was identified for hyaline cartilage versus age, and that further investigation is necessary to better understand and confirm the relationship between hyaline cartilage and age in FAIS. In addition to age, correlations between tissue features and biological sex were also assessed. It is also important to consider the sex-based differences in the rates of skeletal maturity, which may imply that these factors may interact with each other to affect the tissue environment.

### Limitations

In this study, only representative samples of the cam deformity and capsule were retrieved, which may not capture all features that are present. These data were also collected without context of the patient’s clinical presentation. To mitigate this limitation, we strived to obtain samples at the highest point of the cam deformity, to improve consistency between procurements. Despite this, our findings provide a robust quantitative evaluation of tissue features in patients with cam-type FAIS, serving as a major step towards understanding the tissue environment associated with FAIS.

Another potential limitation of the presented study is the lack of a control group for comparison. However, it is crucial to note that in this study, it would be unethical to remove healthy normal tissue from either a patient with FAI or a healthy individual, as this may incur harm on human subject. While one could argue that cadavers may be used as a “control” specimen, we chose not to use cadaveric specimens, as they are often fresh frozen, which can cause artifacts in histological preparation. Therefore, a one-to-one comparison between surgically retrieved pathological samples and cadaveric samples would not be a reasonable comparison. Therefore, the purpose of this study was not to compare FAI vs. healthy groups, but to comprehensively assess the overall variability of tissue structures in the cam deformity, to better understand how consistent the cam deformity’s tissue structure is in the context of FAI.

## CONCLUSION

The cam deformity is complex and heterogeneous, both within individual cam deformities and between individuals with FAIS. Several cam deformity tissue features were correlated with α angle, age, sex, and BMI. The heterogeneity observed in these samples indicates that tissue properties within the cam deformity varies between patients with FAIS, which may contribute to outcomes of hip arthroscopic surgery and a patient’s level of risk for the subsequent development of osteoarthritis.

## Data Availability

All data produced in the present study are available upon reasonable request to the authors.

## Acknowledgements

The authors would like to thank Dr. Cecilia Pascual-Garrido for guidance on method development and data interpretation.

